# Individual and community-level determinants of intention to use contraceptive among married women in Ethiopia: A multi-level analysis of National Survey

**DOI:** 10.1101/2022.02.07.22270245

**Authors:** Kegnie Shitu, Adugnaw Zeleke Alem, Tesfa Sewunet Alemneh, Bewuketu Terefe

**Affiliations:** Department of Health Education and Behavioral Science, Institute of Public Health, College of Medicine and Health Sciences, University of Gondar, Gondar, Ethiopia; Department of Epidemiology and Biostatistics, Institute of Public Health, College of Medicine and Health Sciences, University of Gondar, Gondar, Ethiopia; Department of Community Health Nursing, School of Nursing, College of Medicine and Health Sciences, University of Gondar, Gondar, Ethiopia

**Keywords:** Intention to contraception, married women, Multilevel analysis, Contraceptive, Ethiopia

## Abstract

**Background:** Contraceptives are the most effective strategies to prevent unwanted pregnancies and their consequences. Realizing intention to use contraceptives is a crucial stage to draft and implement a successful family planning program.

**Objective:** This study aimed to identify individual and community level of factors affecting women’s intention to use contraceptives in Ethiopia.

**Method:** This study was based on a large national survey, Ethiopian Demographic and Health Survey. A total weighted sample of 6,555 married/in union reproductive-age women were included. Because of the hierarchical nature of the DHS data, a multilevel logistic regression model was used to study individual and community-level factors that may influence intention to contraceptive use. A 95% confidence interval and a p value of less than 0.05 were used to declare statistical significance.

**Result:** The overall intention to use contraceptives was 48.63% (95% CI 47.42, 49.84). Participants’ age range of 25-34 years (AOR = 0.42, 95 CI% (0.22, 0.79)) and 35-49 years (AOR = 0.12, 95% CI: (0.05, 0.28)), husband with primary education (AOR = 1.60, 95% CI: (1.02, 2.50)), heard of contraceptives from their community (AOR = 1.91. 95% CI: (1.29, 2.83)), ever used contraceptives (AOR = 4.48, 95% CI: (2.91, 6.88)) and having six or more children (AOR = 0.46, 95% CI: (0.23, 0.9)) were individual factors significantly associated with interceptive intention. From community level factors, high community family planning utilization rate (AOR = 2.29, 95% CI: (1.36,3.86)) was associated with intention to use contraceptive.

**Conclusion:** More than half of married women were not intended to use contraceptives. Intention to use contraceptive was affected by individual and community level attributes. Thus, public health interventions particularly that could increase information dissemination regarding contraceptives among the communities and enhance community level contraceptive utilization rate are required at the national level to improve contraceptive utilization.

## Introduction

A couple’s purposeful effort to restrict or space the number of children they have through the use of contraceptive techniques is referred to as family planning. Contraceptive methods are divided into two categories: modern and old. Male and female sterilization, the intrauterine contraceptive device (IUD), implants, injectables, the pill, male and female condoms, emergency contraception, the standard 9 days method (SDM), and the lactational amenorrhea method are all examples of modern approaches (LAM). Traditional approaches include things like rhythm, withdrawal, and other folk methods(1).

Reports pointed out that, in 2019 there were about 1.1 billion reproductive age women who have a need to use contraceptive methods, however, around 270 million of them had faced unmet need of contraceptive methods(2, 3). Sustainable development goals (SDG) and united nations world family planning report indicated that the total of rate satisfaction of mothers in family planning is stopped at around 78%, nevertheless due to several hindering factors the rate satisfaction in Africa regions has shown lowest rate (56%)(4). If all low and middle countries can satisfy women in contraceptive services, they will have a chance of reduced unintended pregnancies, unplanned births, and induced abortions approximately by three-quarters from 39 million to 22 million, from 30 million to 7 million and from 48 million to 12 million every single year respectively with 76,000 fewer maternal deaths(5).

In the present scientific fact, the global community has reached on consensus agreements regarding family planning by reducing the number of women’s exposure to pregnancies, by limited more than 4 births (6, 7), reducing unsafe abortion from unintended pregnancies (8, 9), declining newborn and infant mortality rates (5), preventing the transmission of HIV/AIDS from the mother to the child by 93% (10), empowering mothers and girls by staying them in school(11), by improving adolescents’ reproductive health, social and economic wellbeing(12). The task of family planning is exponentially difficult and daunting in sub Saharan Africa (SSA), hence According to the world bank project report the number of populations SSA has shown an increasement of about ten times between 1990 and 2050(13), this in turn shows population growth is fast in Africa(14). SSA has shown a few declines from 6.3 to 4.6 births per women from 1990 to 2019(14).

Like the rest of the world community, Ethiopia is struggling with family planning issues and has tried to increase contraception coverage by 69% by 2020, however has failed and it needs other fertility and family planning implications (15). Studies on contraception prevalence in Ethiopia show that it is low and far apart in terms of prevalence from one to other studies. For illustrations, a study conducted in Benishangul Gumuz revealed that the prevalence of contraceptive was 18.6%(16), 38% in southern Ethiopia Wolayita Sodo(17), 19.9% in eastern Ethiopia (18). Other two important studies were also conducted based on demographic and health survey data on contraceptive prevalence to use and discontinuation discovered that 20.42% and 32.2% respectively(19, 20). Children born from mothers who use modern contraceptive methods have high rate of survival implications than their counterparts(21). Such studies also identified some crucial factors which have positive or negative statistical implications on contraceptive method use. This includes age, educational level, knowledge, attitude (16, 17), residence, region, occupational status, number of children, women self-decision, television watching (19, 20), counseling on contraceptive methods, (16, 20), myths and misconceptions(17). There were also some other factors associated with intention to use contraceptive methods. Fear of side effects, male partner opposition, inter spousal discussion, discussion with health care providers, perceived cultural acceptability, status of gravidity, marital status and post-natal care utilization were among the variables associated with the outcome variable(22–27).

This study addresses two major research objectives. First, studies done on contraceptive methods in Ethiopia are so limited in study area and study population that is difficult to determine for the general population of the country. Second, these studies did not include events that may have a potential effect on intention to use contraception methods at community level factors. Therefore, this study aimed to include both individual and community level factors to determine the intention to use contraceptive methods among married reproductive women. Identifying important factors affecting intention to use contraceptive methods at the community and individual level, this study contributes to Ethiopia, would have a paramount importance to increase birth control by controlling birth rates. Policymakers and other stakeholders working on family planning, women and children’s health will access an up-to-date nationwide result from the findings of this study to develop, improve and implement their plans accordingly.

## Methods

### Data source, population, and sampling procedure

The present study was based on the most recent Ethiopian Demographic and Health Survey (EDHS) data. The survey was conducted from January 18, 2016, to June 27, 2016. For this survey, a complete list of 84, 915 Enumeration Areas (EAs) from the Ethiopian Population and Housing Census was used as a sampling frame. A stratified two-stage cluster sampling technique was employed. In the first stage, 645 EAs were selected. In the second stage, an average of 28 households was selected per cluster/EA. The data is freely available in public and we accessed the dataset used for the present study after we registered and received an authentication letter from the Demographic and Health Survey (DHS) program at The DHS Program - Ethiopia: Standard DHS, 2016 Dataset. For this study, a total weighted sample of 6, 555 married/in union women who are not using contraceptive were included. Detailed information about the sampling strategy, questioner, or other important information is available in EDHS of 2016(28).

### Variables of the Study

The outcome variable of the present study was the intention to use contraceptives among contraceptive non-user women. The variable was dichotomized into **1** = “Intended” and **0** = “Not intended”. The independent variables were further classified into individual level (level 1) variables and community level (level 2) variables. Individual-level variables included age, religion, educational status, husband’s/partner’s age and education, family wealth index, current working status, family planning message exposure, knowledge of family planning methods, ever use of contraceptive, fertility preference, desire to have children, and a number of children. Whereas, community variables involved variables directly taken with no aggregation (residence and contextual region), and variables obtained by aggregating individual variables into their respected community (community poverty, community female education, community family planning message exposure, community antenatal care service utilization rate, community family planning utilization rate, community women’s education, and community health facility distance). The aggregates were computed using the mean values of the proportions of women in each category of a given variable. Since the aggregate values of each variable don’t follow a normal distribution curve, we categorized the aggregate values of a cluster into groups based on median values.

### Operational definitions

#### Household wealth quintile

The wealth index classifications were done in quintiles: poorest; poor; middle; rich; richest. These were computed using principal component analyses (PCA). This variable was further categorized into three categories (Poor, Medium, and Rich) by merging the lower two (poorest and poor) quantiles and the upper two (richest and rich) quantiles.

#### Family planning message exposure

This variable was computed from the frequency of exposure to family planning messages from television, radio, magazine or mobile messages The variable was categorized into two parts: Not exposed when the participant is exposed to none of the four channels at least once a week; and exposed if the participant was exposed to family planning messages at least once a week from one of the four channels.

#### Contextual region

For this survey regions were categorized into three categories (agrarian, pastoral, and metropolitan) that may have a strong relationship to health information seeking and intention to use contraceptives. The Tigray, Amhara, Oromia, SNNP, Gambella, and Beneshangul Gumuz were recorded as agrarian. The Somali and Afar regions were merged to form the pastoralist region and the city administrations-Addis Ababa, Dire Dawa, and Harar were combined as metropolitan (1).

#### Community female education

This is the aggregate value of the educational levels of women based on the average of proportions of educational levels in the community. It was defined as low if the proportion of women with secondary education & above in the community was 0 –12.4 % and high if the proportion was 12.5 –100 %.

#### Community media exposure

This variable was derived from the individual responses for exposure to radio or television. It was defined as low if the proportion of women exposed to media in the community was 0–18.7 % and high if the proportion was 18.8–100 %.

#### Community ANC utilization rate

This variable is also derived from the individual values for ANC utilization. It was defined as low if the proportion of women who attended at least one ANC visit in the community was 0 – 81.3 % and high if the proportion was between 81.4 –100%.

#### Community poverty

With the same procedure, this variable is also derived from an individual household’s wealth index. It was defined as high if the proportion of women from the two lowest wealth quintiles in a given community was 25.9–100 % and low if the proportion was 0–26 %.

#### Community distance to the health facility

The variable was aggregated from individual perceived distance to a health facility is a big problem. It was as categorized as low if the proportion of women who perceived health facility distance as a big problem in the community was 0–42.2% and categorized as high if the proportion was between 42.2% and 100%.

### Data processing and Analysis

Data were extracted from individual records (IR) files and further coding and transformations were done using statistical software, STATA version 14. The weighted samples were used for analysis to adjust for unequal probability of selection and non-response in the original survey. Since the EDHS employed multi-stage stratified cluster sampling techniques, the data have a hierarchical structure. In such kinds of situations, single-level logistic regression is not recommended because Traditional multiple regression techniques treat the units of analysis as independent observations. One consequence of failing to recognize hierarchical structures is that standard errors of regression coefficients will be underestimated, leading to an overstatement of statistical significance. In this point of view, to draw valid inference and conclusion an advanced statistical model which takes the hierarchy of the data into account is required. Therefore, a multivariable multilevel binary logistic regression model was used to estimate the fixed and random effects of the factors associated with intention to use contraceptives. Four models were constructed. The first model also called an empty model which was fitted without any explanatory variables. This model was specified to decompose the amount of variance that existed between communities. The null model is important for understanding the community variations, and we used it as a reference to estimate how much community factors were able to explain the observed variations in the intention to use contraceptives. Moreover, this model was used to justify the use of a multilevel statistical framework as it is a litmus paper on whether multilevel or the traditional logistic regression should be used. It was assessed using the Log Likelihood Ratio test (LLR), Median Odds Ratio (MOR), Intra-class Correlation Coefficient (ICC), and Proportional Change of Variance (PCV). The second model contained only individual-level factors. The third contained only community-level factors. Whereas, the final (fourth) model containing both individual and community-level factors. Moreover, the model comparison was done using model deviance, a model with the lowest deviance selected for reporting and interpretation results.

## Result

### Individual level characteristics

A total of 6,555 weighted sample of married/ in union and contraceptive non-users were involved in the present study. The median age of the women was 30 years old with an interquartile range (IQR) of 13 years. The median (IQR) husband’s/partner’s age was 38 (16) years. More than half of (56.6%) of the participants were affiliated to Christianity. Majority (65.68%) and more than half (50.51%) of the participants and their husbands/partners did not attend formal education respectively. About 2999 (46%) of the participant were form a house hold with poor wealth status. Concerning exposure to family planning messages, only 1572 (24%) of the participants were exposed to family planning message from at least one of the four main (television, radio, magazine and mobile message) channels.

Almost all (98%) of the participants knew at least one of the family planning methods. However, only 2257 (34.4%) of the participants had ever used at least once a time. Decision for not using contraceptive was mainly made jointly by the respondents and their partner whereas 569 (10.43) participants reported that their partners were the main decision makers for not using contraceptives. Regarding to fertility, about 2840 (43.3%) participants hadn’t desire to have children in the future. In more than on third (35.3%) the cases both the respondents and their partners want the same number of children to have in the future (Table 1).

**Table 1:**
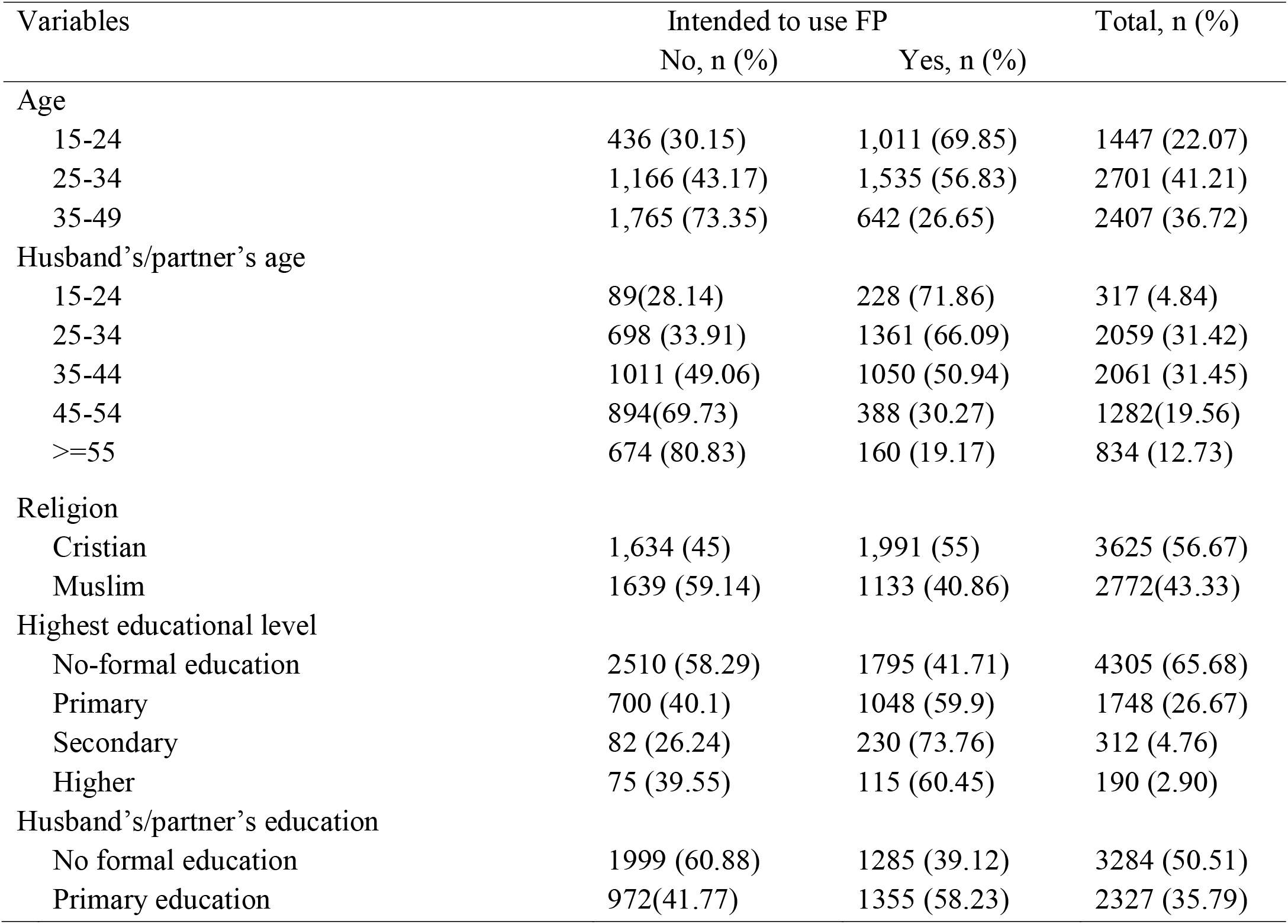
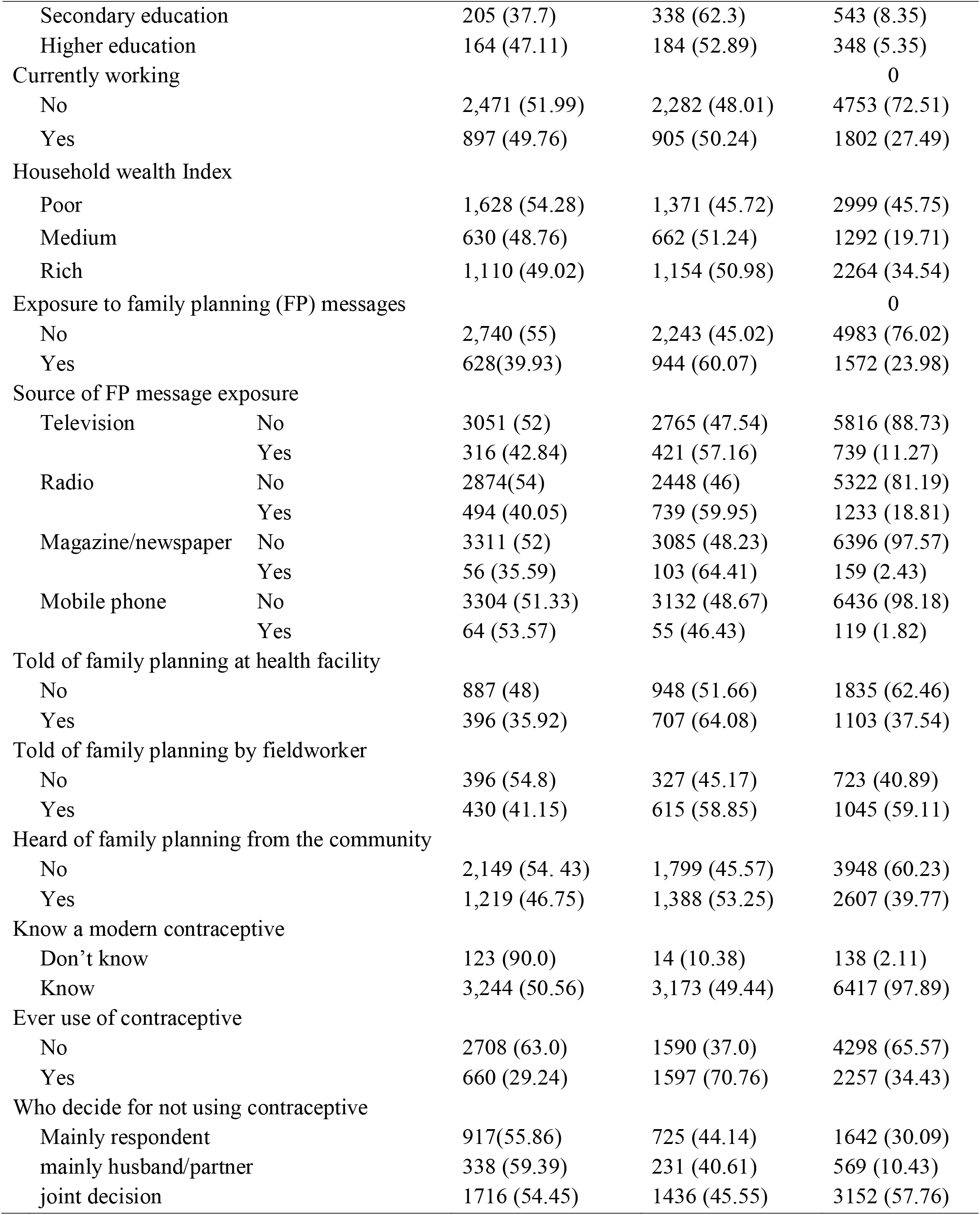
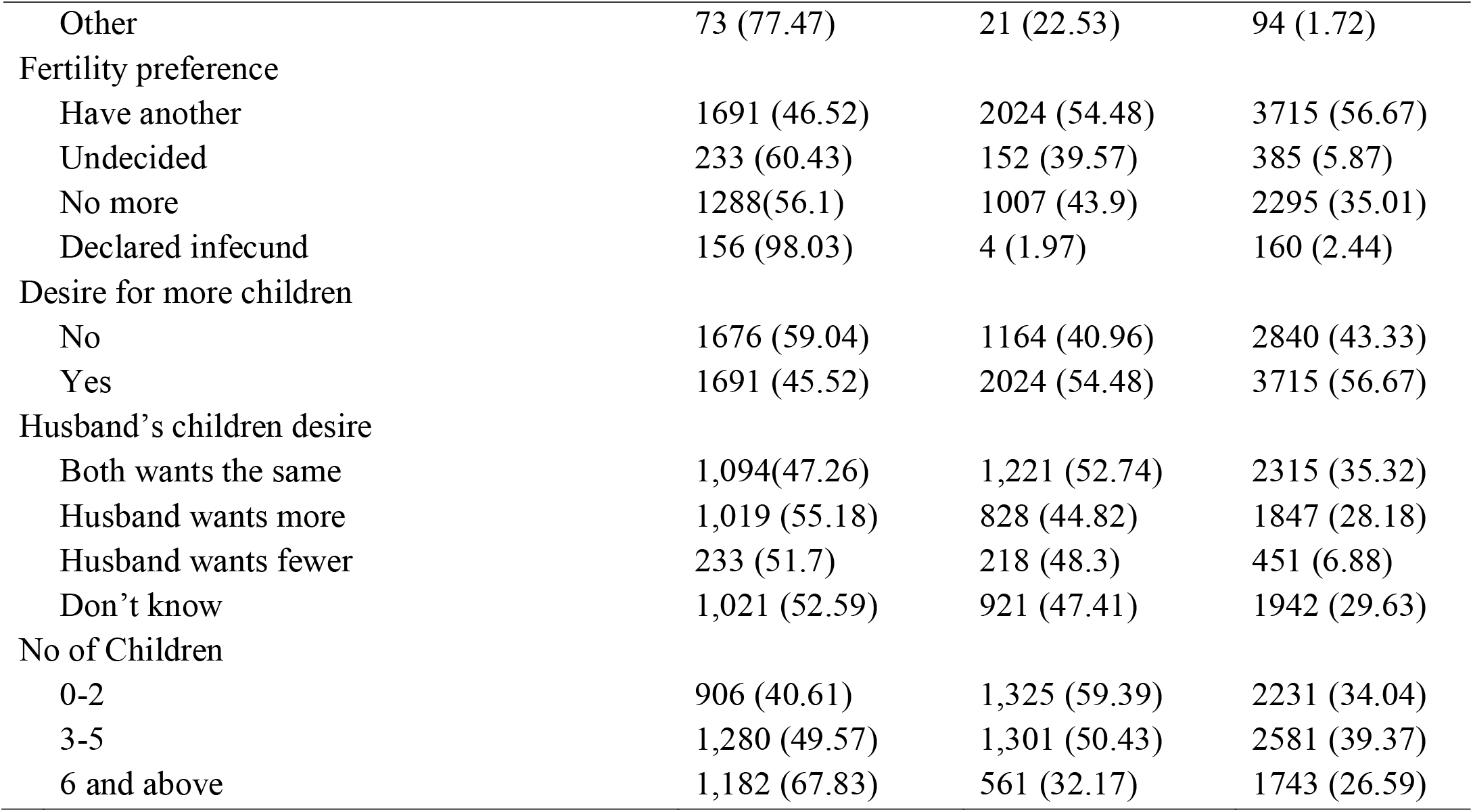
Sociodemographic and other individual characteristics of married/in union women in Ethiopia (n=6555)

| Variables | Intended to use FP |  | Total, n (%) |
| --- | --- | --- | --- |
|  | No, n (%) | Yes, n (%) |  |
| Age |  |  |  |
| 15-24 | 436 (30.15) | 1,011 (69.85) | 1447 (22.07) |
| 25-34 | 1,166 (43.17) | 1,535 (56.83) | 2701 (41.21) |
| 35-49 | 1,765 (73.35) | 642 (26.65) | 2407 (36.72) |
| Husband's/partner's age |  |  |  |
| 15-24 | 89(28.14) | 228 (71.86) | 317 (4.84) |
| 25-34 | 698 (33.91) | 1361 (66.09) | 2059 (31.42) |
| 35-44 | 1011 (49.06) | 1050 (50.94) | 2061 (31.45) |
| 45-54 | 894(69.73) | 388 (30.27) | 1282(19.56) |
| >=55 | 674 (80.83) | 160 (19.17) | 834 (12.73) |
| Religion |  |  |  |
| Cristian | 1,634 (45) | 1,991 (55) | 3625 (56.67) |
| Muslim | 1639 (59.14) | 1133 (40.86) | 2772(43.33) |
| Highest educational level |  |  |  |
| No-formal education | 2510 (58.29) | 1795 (41.71) | 4305 (65.68) |
| Primary | 700 (40.1) | 1048 (59.9) | 1748 (26.67) |
| Secondary | 82 (26.24) | 230 (73.76) | 312 (4.76) |
| Higher | 75 (39.55) | 115 (60.45) | 190 (2.90) |
| Husband's/partner's education |  |  |  |
| No formal education | 1999 (60.88) | 1285 (39.12) | 3284 (50.51) |
| Primary education | 972(41.77) | 1355 (58.23) | 2327 (35.79) |

|  |  |  |  |  |
| --- | --- | --- | --- | --- |
| Secondary education |  | 205 (37.7) | 338 (62.3) | 543 (8.35) |
| Higher education |  | 164 (47.11) | 184 (52.89) | 348 (5.35) |
| Currently working |  |  |  | 0 |
| No |  | 2,471 (51.99) | 2,282 (48.01) | 4753 (72.51) |
| Yes |  | 897 (49.76) | 905 (50.24) | 1802 (27.49) |
| Household wealth Index |  |  |  |  |
| Poor |  | 1,628 (54.28) | 1,371 (45.72) | 2999 (45.75) |
| Medium |  | 630 (48.76) | 662 (51.24) | 1292 (19.71) |
| Rich |  | 1,110 (49.02) | 1,154 (50.98) | 2264 (34.54) |
| Exposure to family planning (FP) messages |  |  |  | 0 |
| No |  | 2,740 (55) | 2,243 (45.02) | 4983 (76.02) |
| Yes |  | 628(39.93) | 944 (60.07) | 1572 (23.98) |
| Source of FP message exposure |  |  |  |  |
| Television | No | 3051 (52) | 2765 (47.54) | 5816 (88.73) |
|  | Yes | 316 (42.84) | 421 (57.16) | 739 (11.27) |
| Radio | No | 2874(54) | 2448 (46) | 5322 (81.19) |
|  | Yes | 494 (40.05) | 739 (59.95) | 1233 (18.81) |
| Magazine/newspaper | No | 3311 (52) | 3085 (48.23) | 6396 (97.57) |
|  | Yes | 56 (35.59) | 103 (64.41) | 159 (2.43) |
| Mobile phone | No | 3304 (51.33) | 3132 (48.67) | 6436 (98.18) |
|  | Yes | 64 (53.57) | 55 (46.43) | 119 (1.82) |
| Told of family planning at health facility |  |  |  |  |
| No |  | 887 (48) | 948 (51.66) | 1835 (62.46) |
| Yes |  | 396 (35.92) | 707 (64.08) | 1103 (37.54) |
| Told of family planning by fieldworker |  |  |  |  |
| No |  | 396 (54.8) | 327 (45.17) | 723 (40.89) |
| Yes |  | 430 (41.15) | 615 (58.85) | 1045 (59.11) |
| Heard of family planning from the community |  |  |  |  |
| No |  | 2,149 (54. 43) | 1,799 (45.57) | 3948 (60.23) |
| Yes |  | 1,219 (46.75) | 1,388 (53.25) | 2607 (39.77) |
| Know a modern contraceptive |  |  |  |  |
| Don't know |  | 123 (90.0) | 14 (10.38) | 138 (2.11) |
| Know |  | 3,244 (50.56) | 3,173 (49.44) | 6417 (97.89) |
| Ever use of contraceptive |  |  |  |  |
| No |  | 2708 (63.0) | 1590 (37.0) | 4298 (65.57) |
| Yes |  | 660 (29.24) | 1597 (70.76) | 2257 (34.43) |
| Who decide for not using contraceptive |  |  |  |  |
| Mainly respondent |  | 917(55.86) | 725 (44.14) | 1642 (30.09) |
| mainly husband/partner |  | 338 (59.39) | 231 (40.61) | 569 (10.43) |
| joint decision |  | 1716 (54.45) | 1436 (45.55) | 3152 (57.76) |
| Other | 73 (77.47) | 21 (22.53) | 94 (1.72) |
| Fertility preference |  |  |  |
| Have another | 1691 (46.52) | 2024 (54.48) | 3715 (56.67) |
| Undecided | 233 (60.43) | 152 (39.57) | 385 (5.87) |
| No more | 1288(56.1) | 1007 (43.9) | 2295 (35.01) |
| Declared infecund | 156 (98.03) | 4 (1.97) | 160 (2.44) |
| Desire for more children |  |  |  |
| No | 1676 (59.04) | 1164 (40.96) | 2840 (43.33) |
| Yes | 1691 (45.52) | 2024 (54.48) | 3715 (56.67) |
| Husband's children desire |  |  |  |
| Both wants the same | 1,094(47.26) | 1,221 (52.74) | 2315 (35.32) |
| Husband wants more | 1,019 (55.18) | 828 (44.82) | 1847 (28.18) |
| Husband wants fewer | 233 (51.7) | 218 (48.3) | 451 (6.88) |
| Don't know | 1,021 (52.59) | 921 (47.41) | 1942 (29.63) |
| No of Children |  |  |  |
| 0-2 | 906 (40.61) | 1,325 (59.39) | 2231 (34.04) |
| 3-5 | 1,280 (49.57) | 1,301 (50.43) | 2581 (39.37) |
| 6 and above | 1,182 (67.83) | 561 (32.17) | 1743 (26.59) |

### Community level factor

Majority (87.84%) of the participants were from rural residency. Whereas, 4178 (63.7%), 4734 (72.2%), 2803 (42.76), 4808 (73.35) and 3748 (57.18) of the participants were from a community with high poverty, low community female’s education, low exposure to family planning, low community ANC utilization rate and low women’s empowerment respectively. Moreover, in the chi2 analysis, residence, community poverty, region, community level female’s education, community ANC utilization rate, community exposure to family planning messages, community family planning utilization rate and community health facility distance problem were significantly associated with intention to use family planning at p-value <0.05 (Table 2).

**Table 2:** Community level variables descriptive result by informed choice among contraceptive user women in Ethiopia (n= 6,555)

| Variables | Intention to use FP, n (%) | Total, n (%) | p-value |
| --- | --- | --- | --- |

|  | No | Yes |  |  |
| --- | --- | --- | --- | --- |
| Residence |  |  |  |  |
| Urban | 394 (49.50) | 403 (50.50) | 797 (12.16) |  |
| Rural | 2,973 (51.63) | 2,785 (48.37) | 5758 (87.84) | 0.003 |
| Contextual Region |  |  |  |  |
| Agrarian | 2,836 (48.41) | 3,023 (51.59) | 5859 (89.38) |  |
| Pastoral | 425 (84.35) | 79 (15.65) | 504 (7.69) |  |
| Metropolitan | 106 (55.50) | 86 (44.50) | 192 (2.93) | <0.001 |
| Community poverty |  |  |  |  |
| Low | 1,202 (50.59) | 1,175 (49.41) | 2377 (36.26) |  |
| High | 2,165 (51.82) | 2,013 (48.18) | 4178 (63.74) | <0.001 |
| Community female education |  |  |  |  |
| Low | 2468 (52.14) | 2266 (47.86) | 4734 (72.22) |  |
| High | 900 (49.39) | 921 (50.61) | 1821 (27.78) | <0.001 |
| Community exposure to FP messages |  |  |  |  |
| Low | 1,498 (53.45) | 1,305 (46.55) | 2803 (42.76) |  |
| High | 1,869 (49.83) | 1,882 (50.17) | 3751 (57.22) | <0.001 |
| Community ANC utilization rate |  |  |  |  |
| Low | 2558 (53.21) | 2250 (46.79) | 4808 (73.35) |  |
| High | 809 (46.33) | 938 (53.67) | 1747 (26.65) | <0.001 |
| Community FP utilization rate |  |  |  |  |
| Low | 1,401 (64.27) | 780 (35.73) | 2181 (33.27) |  |
| High | 1,966 (44.95) | 2,408 (55.05) | 4374 (66.73) | <0.001 |
| Community health facility distance problem |  |  |  |  |
| Low | 996 (48.86) | 1,043 (51.14) | 2039 (31.11) |  |
| High | 2,371 (52.51) | 2,145 (47.49) | 4516 (68.89) | <0.001 |
| Community women's empowerment |  |  |  |  |
| Low | 1,979 (52.82) | 1,769 (47.18) | 3748 (57.18) |  |
| High | 1,388 (49.45) | 1,419 (50.55) | 2807 (42.82) | 0.287 |

*Not: ANC= Antenatal care, FP= Family planning*

**Table 3:**
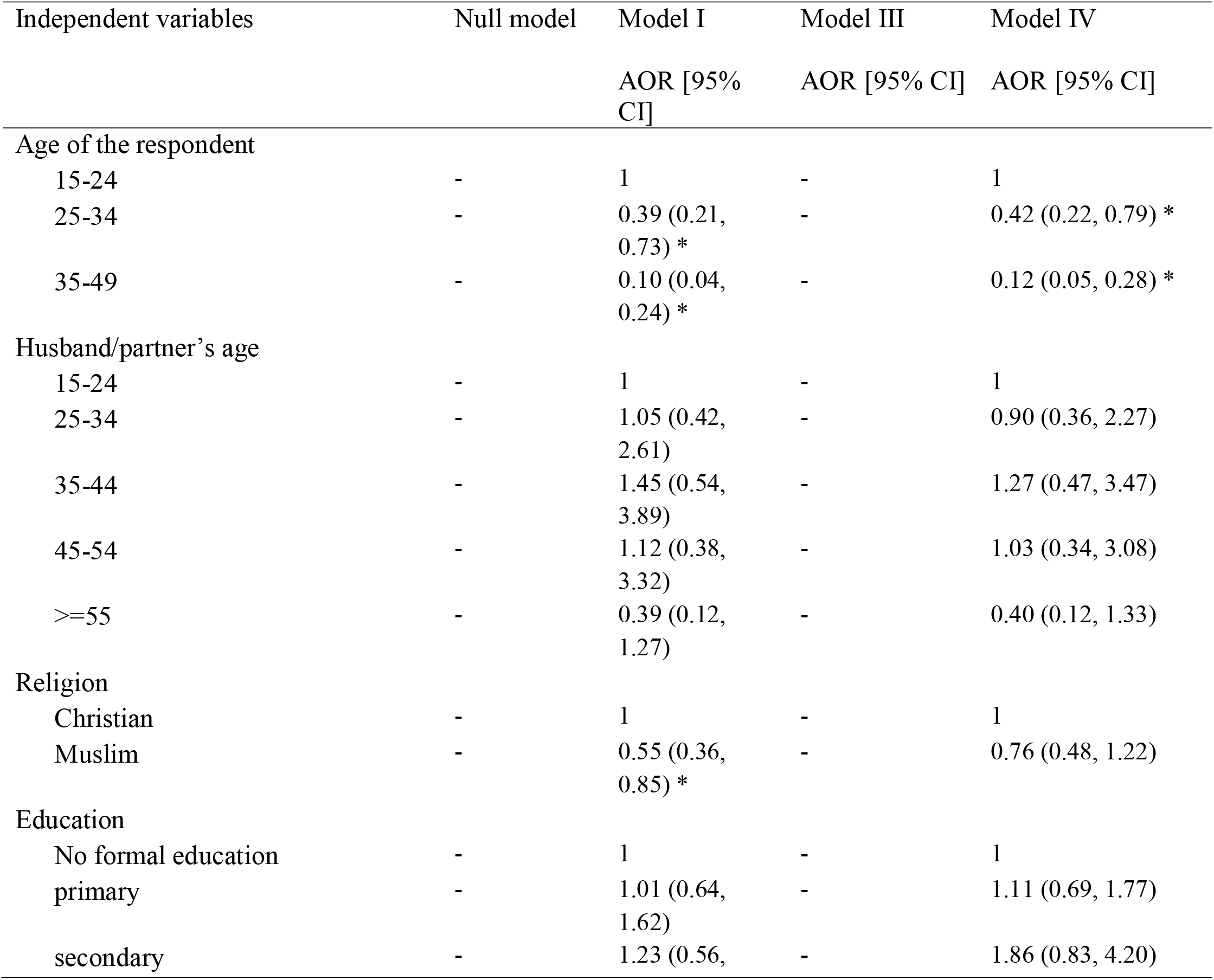
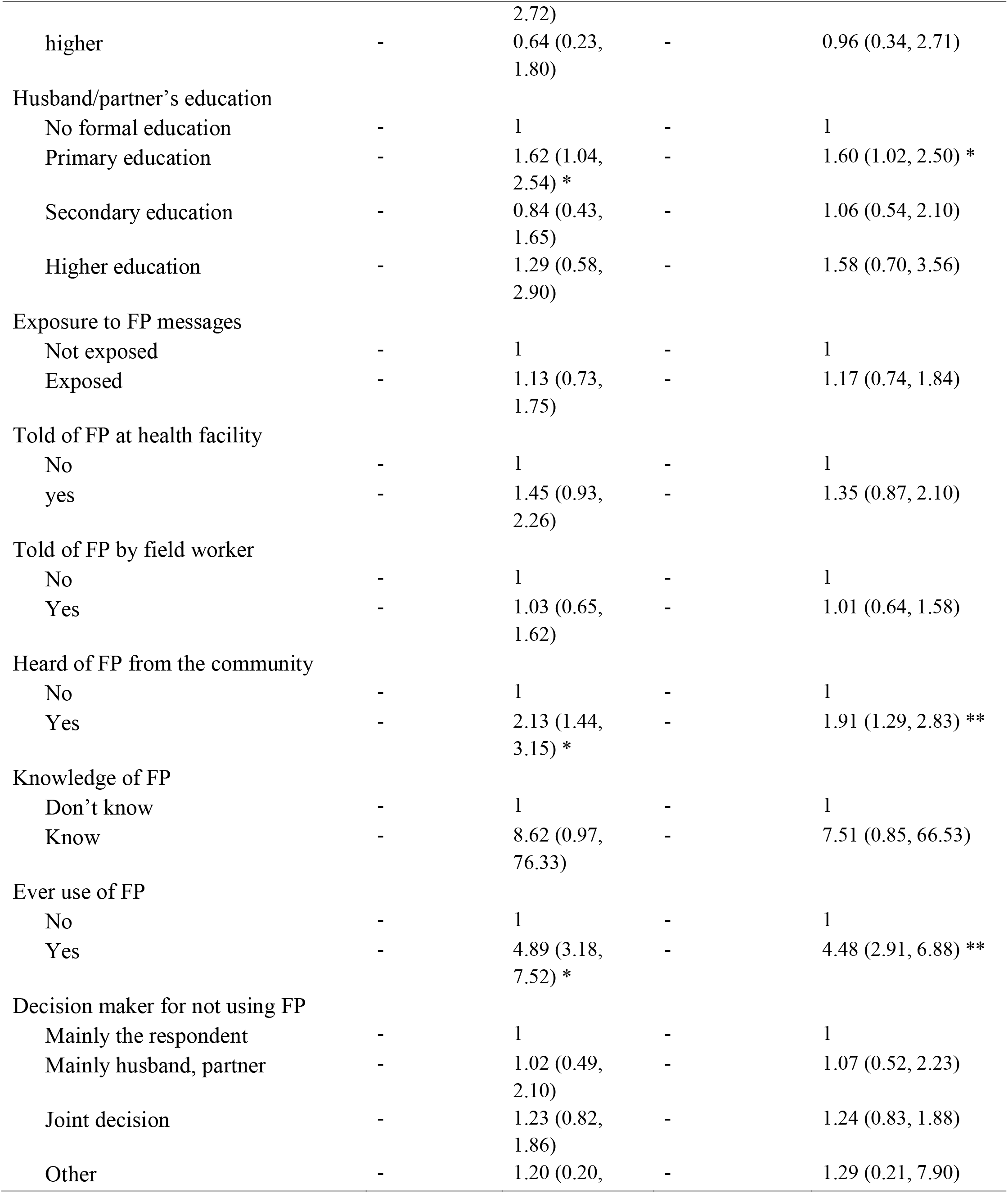
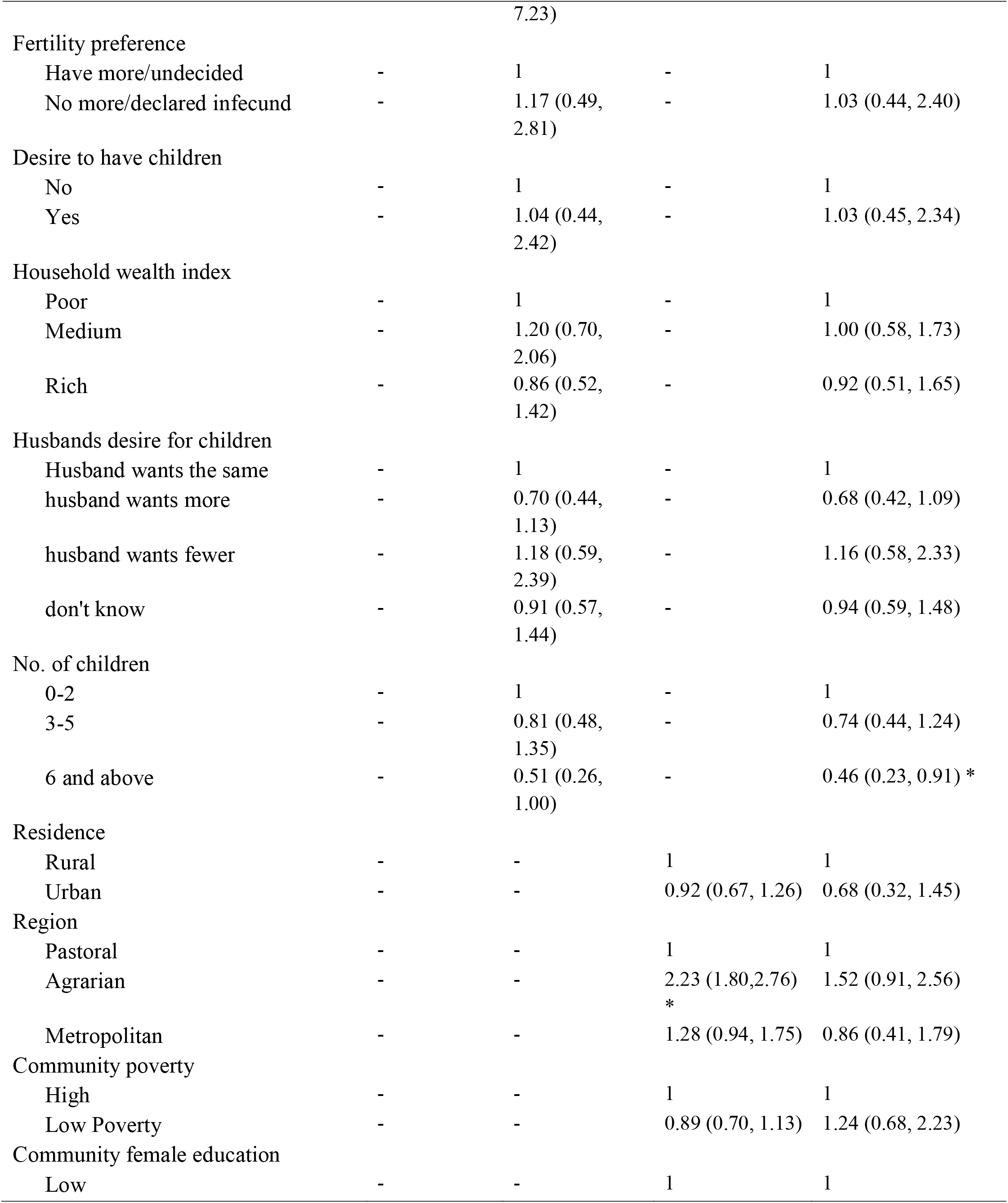
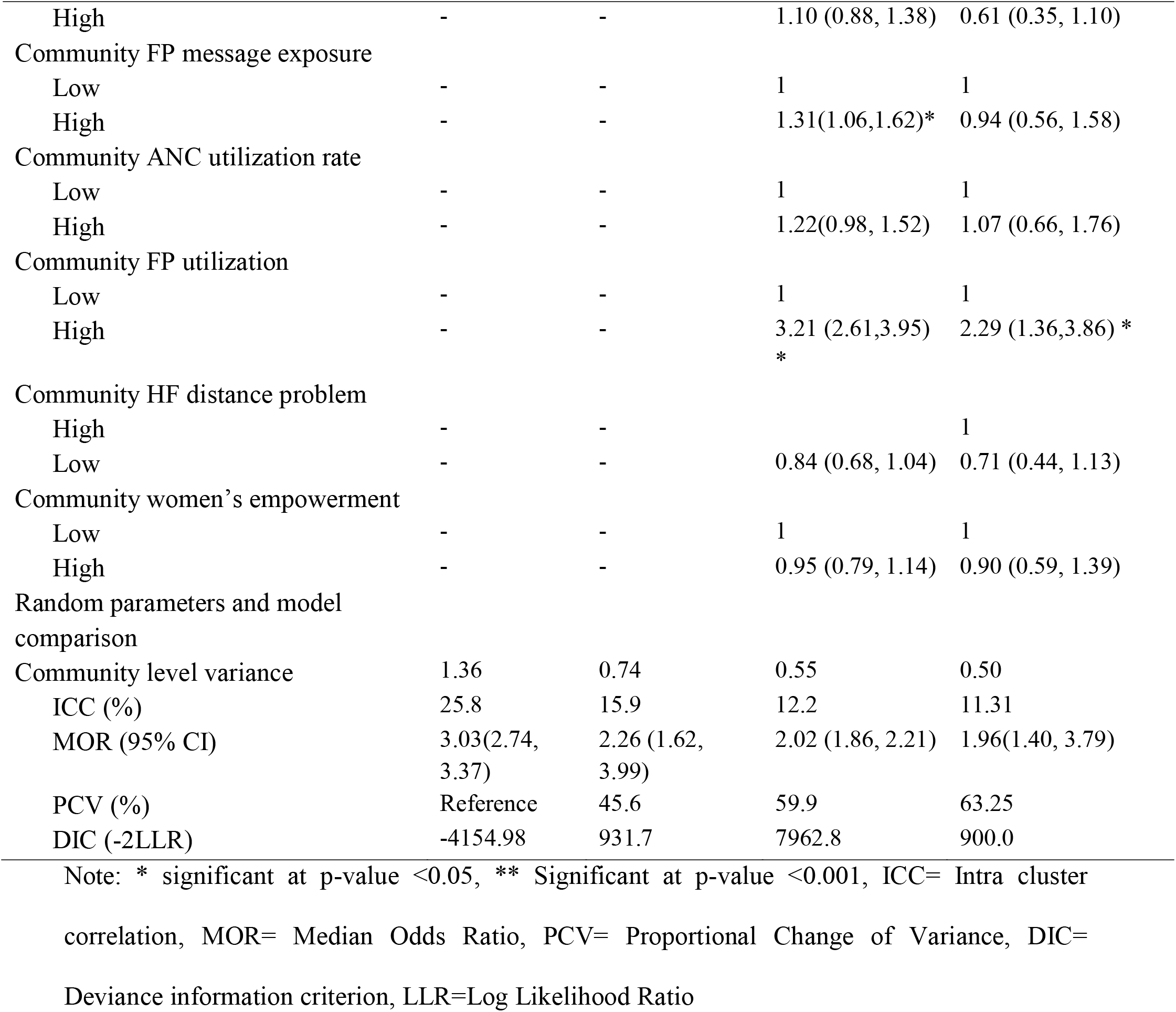
Individual and community level facto associated with intention to use family planning among married/ in union contraceptive non-user women in Ethiopia (n=6,555).

## Intention to use contraceptive

Only 3368 (48.6%, 95% CI 47.42, 49.84) of married reproductive age women were intended to use contraceptive in the future (Figure 1).

**Figure 1:**
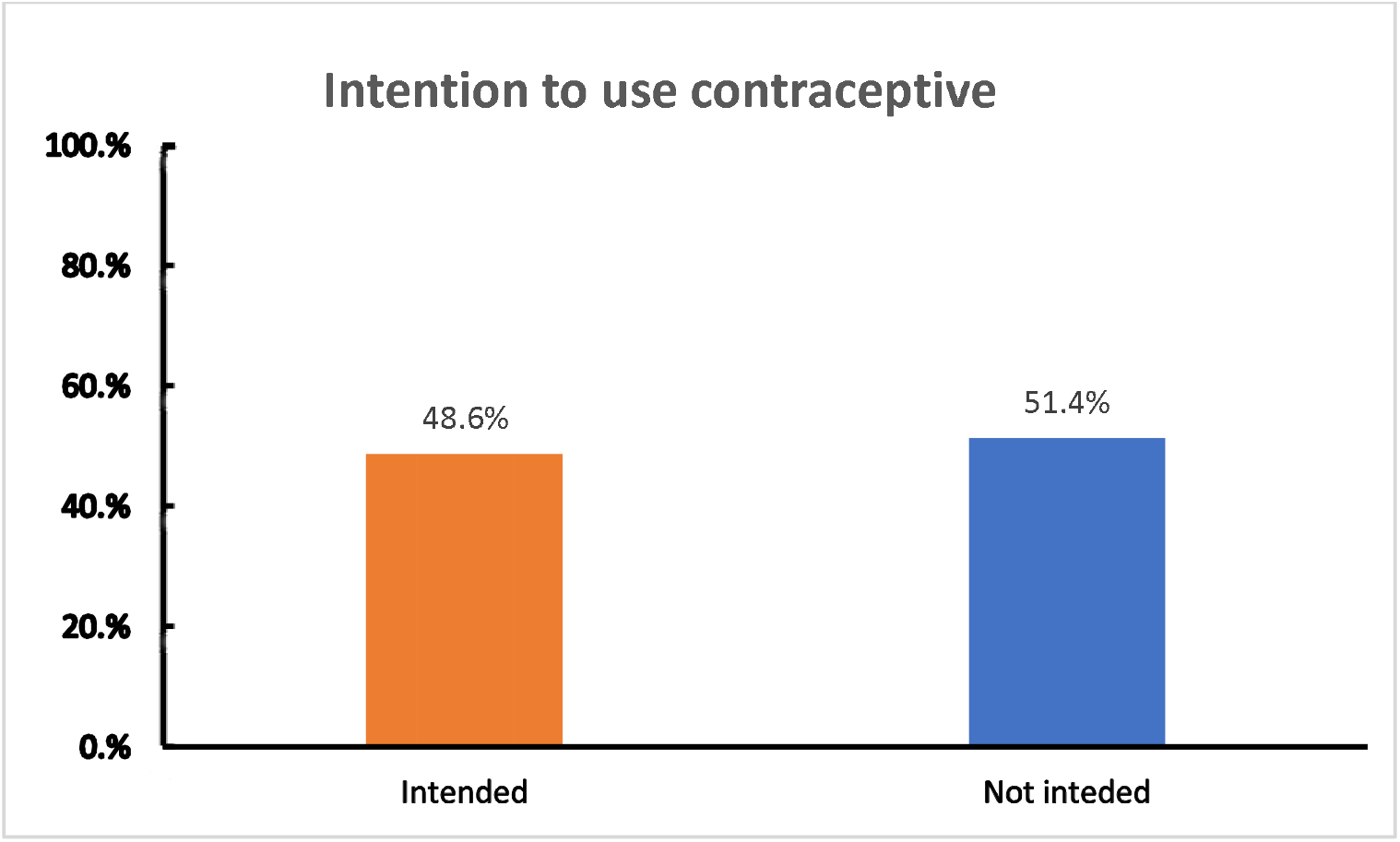
Intention to use contraceptives among married reproductive age women in Ethiopia (n= 6, 555)

### Factors associated with intention to use family planning

In the multilevel multivariable analysis, age, education, heard of family planning from th community, ever use of contraceptives, number of children, and community level family planning utilization rate were significant factors associated with participant’s intention to us contraceptives in the future.

Participants within the age range of 25-34 years (AOR = 0.42, 95 CI% (0.22, 0.79)) and 35-49 years (AOR = 0.12, 95% CI: (0.05, 0.28)) were less likely to be intended to use contraceptives in the future compared to participants with the age range of 15-24 years. Women’s husband/partner with primary education were 1.6 times more likely to be intended to use contraceptives compared to husbands with no formal education (AOR = 1.60, 95% CI: (1.02, 2.50)). Participants heard of contraceptives from their community (AOR = 1.91. 95% CI: (1.29, 2.83)) and ever used contraceptives (AOR = 4.48, 95% CI: (2.91, 6.88)) were almost two and 4.5 time more likely to be intended to use contraceptives in the future respectively. In addition to this, women who had six or more children (AOR = 0.46, 95% CI: (0.23, 0.9)) were less likely to be intended to use contraceptives. Regarding community level variables, Women from a community with high rate of family planning utilization were 2.3 times more likely to be intended to use contraceptives (AOR = 2.29, 95% CI: (1.36,3.86)).

## Discussion

The aim of this study was to assess future intention to use contraceptives and associated factors among married/ in union women who were not current contraceptive user. The proportion of participants who were intended to use contraceptive in the future was 48.63 (47.42-49.84). This figure is less observed than studies conducted in Indonesia 63%(29), in Ghana 70%(30, 31), in Uganda(32), in Malawi (33), in Ethiopia 84.3% (34) and it is almost in agreement with studies conducted in Ghana 49.3%(35), however this figure is higher than studies investigated in Ethiopia (17, 36). The possible justification why the observed findings of this study is less than the above mentioned one might be due to countries profiles of family planning experience, participants attitude, knowledge and education backgrounds towards family planning intention. On the other hand, this study founds significantly higher prevalence of intention to use contraceptives, might be regarding to minimizing confounding factors both on the individuals and community level factors that could have positive or negative implication on the intention to use contraceptives. Majority of those studies concluded the intention to use contraceptive based on individual factors with a very limited sample size and setting of the study. This might have increased their prevalence findings.

This study discovered that age was one of the main predictor’s variables. Participants whose age group is from 25 to 34 years and 35 to 49 years old have less likely intention to use contraceptives in their future family than the youngers. This finding is in agreement with many other studies conducted at various study setting with different times. A study conducted in Cameron (37), in Zambia(38), in Uganda (32), in Malawi (33), and in Ethiopia (39). This is related to the fact that older women may be in less need of contraceptive methods(40). This also can be explained by as mothers gets older and older, their ability to give birth may decrease and they may experience health problems, sometimes due to pregnancy or related reasons. On the other hand, they may think they are infertile due to their age and also may have a negative view of modern contraceptives.

The study also depicted that those women’s husband who have formal education has more likely intention to use contraceptive in the near future than their counterparts having no formal education. This investigation almost similar with other studies done in Ethiopia (23, 41). In Pakistan (42), in Cameroon (43), in Uttar Pradesh (44) and in Nigeria (45). Male partners, especially in rural Ethiopia, have a tradition of domination family matters, sometimes by their own interests. Education is the major key to solve this challenges of self-determination in family issues (46, 47). Education can enhance humans romantic relationship, sense of humor and ability to make informed decision making cultures with their female partners (48, 49). The results of this study also illustrate this fact.

The other interesting variable was those participants who had heard from the community about contraceptive or family planning have more tendency of intention to use contraceptives. This finding is supported by several studies conducted in low and middle income countries’ (50), in Ethiopia(51), in SSA (52), in Rwanda(53), and in Nigeria (54). This shows that a favorable community level information of contraceptives were significant predictors of intention to use modern contraceptive methods and should take into consideration during the development of Ethiopian family planning programs. The increase in the number of people who have heard of the positive benefits of contraceptives in the community is one indication that mothers are ready to do whatever is best for the community.

Individuals who have ever used family planning at least once in their life time has more favorable interest to use the contraceptive methods once again in their future life. These participants who have ever used contraceptives at least once may increase their awareness and knowledge about the service over time. Their awareness and knowledge in turn might increase the intention to use contraceptives through the exposure and counseling received from health care providers. This idea is supported by studies conducted in Ethiopia (34, 39), in Yemen(55), and in Malaysia (56)

Mothers having six and more than six children have shown less positive intention to apply family planning in the near future than their counterparts. This finding is in disagreement studies conducted in Ethiopia (39, 57). The reason why mothers with six or more than six number of children do not use contraceptives is because they might have a low and negative attitude of contraception or have much pressure and future need of children on themselves, their partners or the community to have more children or think they will not have children later due to their aging or uneducated. They may be also using contraceptive method is offensive and forbidden in their religion and cultural practices (58). Those participants might have also faced geographic challenges and unavailability of resources (59). Some wealthier women also mentioned that using contraceptive methods have health concern issues and infrequent sex (50).

Participants living in a community which family planning coverage is more utilized have a greater positive tendency to apply family planning for future upcoming tears than those living in less utilized communities. This finding is supported by studies conducted in Pakistan, Rwanda and Taiwan with a conclusion of accessing of family planning services did not depend only mothers own individual level factors but also by the factors of her household and community level factors (60–62). Other study in Ethiopia also again showed that, Ethiopia will more privilege through community based outreach, and interpersonal communication that could effectively modified the knowledge and behavior of women of intention to use contraceptives (63). Certain community members are almost similar in the knowledge, attitudes and way of life. Thus, it is important for mothers to decide whether or not to use contraceptives. For example, when people around her who have a positive view of modern contraception and have a better quality and a healthier baby, mother will develop more courage and self-confidence the intention to use modern contraceptives. Perhaps she could explain to others in her community that using contraception would be better for her, her baby and her family. Or, if she has a misconception view of birth control, she is more likely to benefit from it if the majority of the community benefits. She can easily find the information he needs through her friends and community.

## Conclusion

The findings of this study revealed that intention to use contraceptives in the near future among married women current contraceptive nonusers was very low. Age of women, educational status of male partners, ever use of contraceptives, heard of contraceptives information from the community, having six and a greater number of children and higher community utilization family planning utilization rate were factors that statistically significant variables associated with intention to use contraceptives. Thus, public health interventions particularly that could increase information dissemination regarding contraceptives among the communities and enhance community level contraceptive utilization rate are urgently required at the national level to address potential hindering factors and to improve the rate of contraceptive utilization in Ethiopia.

### Strength and limitation of the study

The strength of this study includes the use of a large sample size. As the sample size increases, the sample gets closer to the actual population, which decreases the potential for deviations from the actual population. In addition to this, an advanced statistical model that can take the nature of the data into account was employed. However, the present study should be interpreted with several limitation that includes; because of the crossectional nature of the study, making casual inferences about the observed associations might not be possible. Moreover, social desirability bias might be introduced as far as the data were entirely based on self-reports.

## Data Availability

Data for this study were obtained from the DHS Program with approval from the data archivist.

https://www.dhsmeasure.com

## Declarations

## Acknowledgments

Authors acknowledged the DHS program office to take the ethical consideration of this study

## Statement of authors contribution

KS: Conceptualization, data curation, formal analysis, investigation, methodology, resources, software, validation, visualization, writing – original draft, writing – review & editing. BT, AZA, TSA: Data curation, formal analysis, investigation, methodology, resources, software, validation, visualization, writing – original draft, writing – review & editing. All the authors read and approved the final manuscript.

## Availability of data

Data for this study were obtained from the DHS Program.

## Consents for publication

Not applicable

## Ethical approval and consent to participate

Ethical approval and permission letter was requested online to DHS program at www.dhsprogram.com to access the data for this study and DHS program was grated the permission through email.

